# Comparing and understanding the experiences of depressive symptoms and suicidal attempts in sexual and gender minorities, and heterosexual cisgender people of Nepal: A mixed-methods study

**DOI:** 10.1101/2025.09.02.25334909

**Authors:** Bipsana Shrestha, Manoj Panthi Kanak, Bibek Poudel, Lisasha Poudel, Prabin Karki, Prashamsa Bhandari, Bimal Patel, Subas Bastola, Anil Shrestha, Rojina Basnet, Bishnu Prasad Choulagai

**Author notes:** Corresponding Author: Bipsana Shrestha.

## Abstract

**Background:** Violence in sexual and gender minorities (SGMs) are multifaceted and intertwined which severely affects the mental well-being of SGM individuals.

**Objective:** To compare and understand depressive symptoms and suicidal attempts in sexual and gender minorities and heterosexual cisgender people.

**Methods:** This is an explanatory sequential mixed-method study with 190 participants. Ten in-depth interviews were conducted. Regression and thematic analysis were conducted to analyse quantitative and qualitative data respectively.

**Findings:** In model 1, the mean depressive symptoms of transgender participants were higher by 2.47 units (95% CI: 0.62, 4.31, p=0.009) compared to cisgender while it was higher by 2.47 units (95% CI: 1.14, 3.80, p<0.001) in sexual minorities compared to heterosexuals. In model 2, the mean depressive symptoms of sexual minorities were higher by 1.55 units (95% CI: 0.12, 2.99, p = 0.034) compared to heterosexuals, and it was higher by 2.11 units (95% CI: 0.71, 3.51, p=0.006) in victims of bullying. Participants who had depressive symptoms (PHQ-9 ≥10) were 7.41 times (95% CI: 2.30, 23.9) more likely to attempt suicide. Qualitative findings showed financial issues, stress from exams, relationship issues and lack of support from friends and family as leading factors of depressive symptoms and suicidal attempts.

**Conclusions:** Depression and suicide are more in SGM than in heterosexual cisgender people pertaining to bullying based on sexual orientation and gender identity.

**Clinical implications:** Inclusive environment should be fostered in health facilities and bullying should be avoided in community to improve health-seeking behaviour and to decrease depression and suicide in SGM.

**What is already known on this topic:** There is a dearth of studies related to sexual and gender minority groups in Nepal, however, studies in other parts of the world have revealed that there is an increased risk of suicide and poor mental health in LGBT people. It has been recommended that more studies are necessary on their mental health issues.

**What this study adds:** This study indicates that there is increased risk of depression and suicide in people from SGM community. However, this risk is not just because of who they identify themselves as or because of their particular sexual preference, but this is ultimately more related to history of getting bullied by society based on their SOGI.

**How this study might affect research, practice or policy:** It is necessary to create an accepting environment in health care facilities as well as society to decrease bullying of SGMs which in turn helps them to alleviate mental health issues.

## Background

Sexual and gender minority (SGM) community, although often perceived to be a homogenous group, comprises people of diverse sexual orientation, gender identity and expression, and sex characteristics (SOGIESC) including Lesbian, Gay, Bisexual, Transgender, Intersex plus (LGBTI+). ^1,2^ Nepal is often portrayed as a pioneer of the LGBTI+ rights movement in South Asia with the 2007 landmark decision made by the Supreme Court of Nepal in favor of Blue Diamond Society which enhanced recognition of a non-binary gender based on a person’s self-determination followed by the Constitution of Nepal, 2015.^3^ However, legal-gender recognition for trans, gender-diverse and intersex persons still remains a huge challenge as the administrative procedure is very ill-informed and unnecessarily medicalized requiring a proof of gender-reassignment surgery or a medical recommendation.^4^ This process is discriminatory and violates the human rights, personal autonomy, dignity and freedom of trans, gender-diverse (TGD) and intersex persons.^4^

Instances of violence against SGM individuals are multifaceted and intertwined, ranging from bodily interventions like genital mutilation of intersex children (regarded as corrective practices), to pathologization of one’s identity via conversion therapies to again ‘correct’ sexual orientation and/or gender identity to larger systemic and legal injustices.^5^ These human rights violations has severely affected the mental well-being of SGM individuals. Consequently, they hesitate to access health services or engage with healthcare providers due to fear of stigma and discrimination. Even if they seek services, they are prone to face denial of care, discriminatory attitudes and inappropriate pathologizing in healthcare settings based on their SOGIESC.^6^

According to a 2014 survey, almost one out of four of the Nepalese SGM people were denied services or treatment by a hospital or a health clinic.^7^

Various studies around the world also suggest that the SGM individuals report higher rates of depression, anxiety and other mental health issues in comparison to their counterparts.^8,9^ Sexual minorities were more likely to admit having mental health disorders than the heterosexual participants when analyzing the WHO mental health survey.^10^ Multiple studies suggest that these harmful mental health outcomes are more severe in TGD individuals in comparison to their cisgender counterparts including sexual minorities.^11^ A metanalytical study among TGD children, adolescents and young adults reveals 28% had a history of lifetime suicidal ideation while 14.8% had ever attempted suicide.^12^ The reason behind this heightened adverse mental health outcomes have been accounted to minority stress which occurs due to stigma and discrimination towards SGM individuals in a society which is deeply heteronormative and cisnormative.^13^

The Sustainable Development Goals of 2030 pledges to “leave no one behind” to decrease inequality along with progressing on average, by drawing attention to marginalized people, avoiding discrimination on the basis of “race, colour, sex, language, religion, political or other opinion, national or social origin, property, birth, disability” as well as “other status”.^14^ Although the 2030 goals do not explicitly talk about the SGM community, they can be talked based on ‘sex’ or ‘other status’. Unfortunately, there is a dearth of information regarding any aspect of health of the SGM individuals in Nepal including research focusing on their mental wellbeing.

By comparing and understanding the depressive symptoms and experiences of suicidal attempts in sexual and gender minorities and heterosexual cisgender people this study aims to contribute to the existing knowledge gap regarding the mental health of the Nepalese LGBTI+ community.

## Objectives

- To find out the prevalence of depressive symptoms and suicidal attempts, and their associated factors in sexual and gender minorities and heterosexual cisgender people.
- To understand the experience of depressive symptoms and suicidal attempts in sexual and gender minorities and heterosexual cisgender people.

## Methods

### Study design and settings

We used an explanatory sequential mixed method study involving two distinct phases. First, PHQ-9 was used to assess depressive symptoms and participants were also asked about the presence or absence of history of suicidal attempts and bullying. Then, qualitative interviews with a subsample of patients who reported presence of suicidal attempts and/or depressive symptoms was done to explain and understand their experiences. The study was conducted in Kathmandu valley comprising densely populated and diversified districts of Nepal (Kathmandu, Lalitpur and Bhaktapur).

### Study population

Sexual and gender minority people aged 18 years and above were taken as a study group. Heterosexual cisgender people from nearby location who were 18 years and above were matched by age with study groups and taken as comparison group.

### Sampling and sample size

Quantitative: A mix of convenience and snowball sampling was used to approach SGMs with the help of INGO/NGOs and CBOs who were working for SGM, while heterosexual cisgender counterparts were taken conveniently from nearby localities of Kathmandu valley. Sensitization to the topic of sexual orientation and gender identity for heterosexual cisgender people was assured before data collection. Sample size (n) 95 each was taken for study group and comparison group making total sample size of 190.

Qualitative: Participants with history of suicidal attempts in last 6 months and/or depressive symptoms in last 2 weeks were recruited purposively. 5 in-depth interviews (IDIs) from each group (10 in total) were taken, sample size decided based on the research reported previously.^27^ Data saturation was achieved from 10 IDIs.

### Tools and techniques

Quantitative: Data was collected via interview method in person or email or phone call, whichever was appropriate on a case-by-case basis by using Kobo toolbox. The Patient Health Questionnaire – PHQ-9 was used for measuring depressive symptoms. It is a likert scale rating from 0 (not present at all) to 3 (almost every day) (total score 27). The cut-off point taken for determining the presence of depressive symptoms was ≥10.^15^ The history of suicidal attempts in past six months and during the entire lifetime was assessed through a yes/no question based on the participant’s memory. Face validity was maintained by pretesting the questionnaire among 10% of the sample size in a similar population. Cronbach’s alpha (0.81) was calculated from the data of pretesting.

Qualitative: IDIs were conducted using a semi-structured interview guideline which was developed based on previous literature.^31,32^. Non-verbal communication was noted down during the time of the interview. Interviews were recorded with consent of the participants.

### Data management and analysis

Quantitative: Data was directly imported from Kobotoolbox into STATA for further analysis. Linear and logistic regressions were done to find out association between dependent and independent variables. Linearity was checked through scatter plot of residuals and fitted values of dependent variables and verified through lowess smoother. F-statistics was found statistically significant (p<0.001). Normality was checked through histogram and Q-Q plot of residuals. Data was not found to be accurately normal. Homoscedasticity was checked through Breusch-Pagan test and unequal variance was observed. To address the non-normality and heteroscedasticity, robust standard error was used in linear regression. Following the variance inflation factors (mean VIF 1.53 and highest VIF 2.18), multicollinearity was also assessed and not observed among independent variables.

Qualitative: All audio-recorded in-depth interviews were transcribed in the Nepali language. The transcripts were analyzed using dedoose software. Thematic analysis was performed. Interesting and significant verbatim were also reported.

## Results

### Quantitative findings

The mean age of participants was 27.92 years with a standard deviation of 7.49. Most (61.6%) of them were heterosexual and most (59.5%) of them identified themselves as cisgender male.

Majority (68.4%) of the participants were single. Only 16.32% of the participants reported a history of mental illness in their family members.

**Table 1.** Sociodemographic characteristics (n = 190)

| **Characteristics** | **Frequency (%)** |
| --- | --- |
| **Age (mean±SD)** | 27.92±7.49 |
| **Sexual orientation** |  |
| Heterosexual | 117 (61.6) |
| Homosexual (Lesbian and Gay) | 46 (24.2) |
| Bisexual | 27 (14.2) |
| **Gender identity** |  |
| Cis-male | 113 (59.5) |
| Cis-female | 55 (29) |
| Trans-male | 5 (2.6) |
| Trans-female | 17 (9) |
| **Education** |  |
| High education | 118 (62.1) |
| Middle education | 63 (33.2) |
| Low education | 9 (4.7) |
| **Marital status** |  |
| Married or living with a partner | 57 (30) |
| Single | 130 (68.4) |
| Divorced/widowed | 3 (1.6) |
| **History of mental illness** |  |
| Present | 31(16.3) |
| Absent | 159 (83.7) |
| **Bullying** |  |
| Present | 43 (22.6) |
| Absent | 147 (77.4) |

10% of the participants had depressive symptoms and 8.4% of the participants admitted that they attempted suicide in their lifetime. Among them, 25% reported attempting suicide in the last six months.

**Table 2.** Prevalence of depressive symptoms and suicidal attempts according to gender identity and sexual orientation (n=190)

| **Variables** | **Groups** |  |  |  |  |
| --- | --- | --- | --- | --- | --- |
|  | **Cisgender** | **Transgender** | **Heterosexual** | **Sexual** | **Total** |
|  | **n(%)** | **n(%)** | **n(%)** | **minorities** | **participants** |
|  |  |  |  | **n(%)** | **n(%)** |
| **Depressive symptoms** |  |  |  |  |  |
| Mean±SD | 4.6±3.8 | 6.1±4.6 | 3.8±3.6 | 6.3±4 | 4.7±3.9 |
| Yes | 16 (9.5) | 3 (13.6) | 5 (4.3) | 14 (19.2) | 19 (10) |
| No | 152 (90.5) | 19 (86.4) | 112 (95.7) | 59 (80.8) | 171 (90) |
| **Suicidal attempts** |  |  |  |  |  |
| Ever | 12 (7.1) | 4 (18.2) | 6 (5.1) | 10 (13.7) | 16 (8.4) |

In model 1, where bullying was not included in the analysis, the participants who were either single or separated or widowed had 1.40 units (95% CI: 0.09, 2.73, p = 0.036) more depressive symptom score on average than those who were married. The mean depressive symptoms score of transgender participants was higher by 2.47 units (95% CI: 0.62, 4.31, p=0.009) compared to cisgender participants. The mean depressive symptoms score of sexual minority participants was higher by 2.47 units (95% CI: 1.14, 3.80, p<0.001) compared to heterosexual participants.

In model 2, where bullying as an independent variable was also included in the analysis, the mean depressive symptoms score of sexual minority participants was higher by 1.55 units (95% CI: 0.12, 2.99, p = 0.034) compared to heterosexual participants. Mean depressive symptoms were higher by 2.11 units (95% CI: 0.71, 3.51, p=0.006) in those who experienced bullying compared to those who had not. Among SGMs, 43.16% reported a history of bullying on the grounds of their SOGI.

No significant interaction was found between gender identity and sexual orientation. Further, F-statistics was found to be significant (p<0.001) implying that at least one of the predictor variables significantly affected the depressive symptoms.

**Table 3.** Factors associated with depressive symptoms (linear regression) (n=190)

| Predictors | Unadjusted |  |  | Model 1 (without bullying) |  |  | Model 2 (with bullying) |  |  |
| --- | --- | --- | --- | --- | --- | --- | --- | --- | --- |
| | $\beta$ | 95%CI | p-value | $\beta$ | 95%CI | p-value | $\beta$ | 95%CI | p-value |
| **Age** | -0.096 | -0.01, -0.18 | **0.021** | -0.07 | -0.15, 0.02 | 0.141 | -0.07 | -0.16, 0.01 | 0.086 |
| **Education** |  |  |  |  |  |  |  |  |  |
| Above class 12 | **Ref** |  |  | **Ref** |  |  | **Ref** |  |  |
| Up to class 12 | 2.27 | 1.15, 3.39 | **<0.001** | 0.99 | -0.32, 2.31 | 0.138 | 1.23 | -0.07, 2.52 | 0.064 |
| **Marital Status** |  |  |  |  |  |  |  |  |  |
| Married | **Ref** |  |  | **Ref** |  |  | **Ref** |  |  |
| Single/separated/widow | 1.89 | 0.68, 3.11 | **0.002** | 1.40 | 0.09, 2.73 | **0.036** | 1.29 | -0.001, 2.58 | 0.05 |
| **Gender identity** |  |  |  |  |  |  |  |  |  |
| Cisgender | **Ref** |  |  | **Ref** |  |  | **Ref** |  |  |
| Transgender | 1.57 | -0.18, 3.34 | 0.079 | 2.47 | 0.62, 4.31 | **0.009** | 1.24 | -0.74, 3.22 | 0.217 |
| **Sexual Orientation** |  |  |  |  |  |  |  |  |  |
| Heterosexual | **Ref** |  |  | **Ref** |  |  | **Ref** |  |  |
| Sexual minorities | 2.51 | 1.40, 3.62 | **<0.001** | 2.47 | 1.14, 3.80 | **<0.001** | 1.55 | 0.12, 2.99 | **0.034** |
| **Bullying** | 3.22 | 1.94, 4.49 | **<0.001** | - | - | - | 2.11 | 0.71, 3.51 | **0.006** |

There was significant association of suicidal attempts with depressive symptoms (p=0.001) in multivariate analysis. Participants who had depressive symptoms (PHQ-9 ≥10) were 7.41 times (95% CI: 2.30, 23.9) more likely to attempt suicide compared to those who did not have depressive symptoms. No interaction effect between gender identity and sexual orientation was found for suicidal attempts as outcome variable.

Additionally, suicidal ideation was found to be significantly associated with depressive symptoms (p<0.001) and with age (p= 0.01), after adjusting for education, gender, sexual orientation and marital status. Participants with depressive symptoms were 5.5 times (95% CI: 2.2, 14.2) more likely to have suicidal ideations compared to participants without depressive symptoms. Likewise, participants with higher age (27-47 years) were 71% (95% CI: 0.11, 0.76) less likely to have suicidal ideations compared to participants with lower age (18-26 years).

**Table 5.**
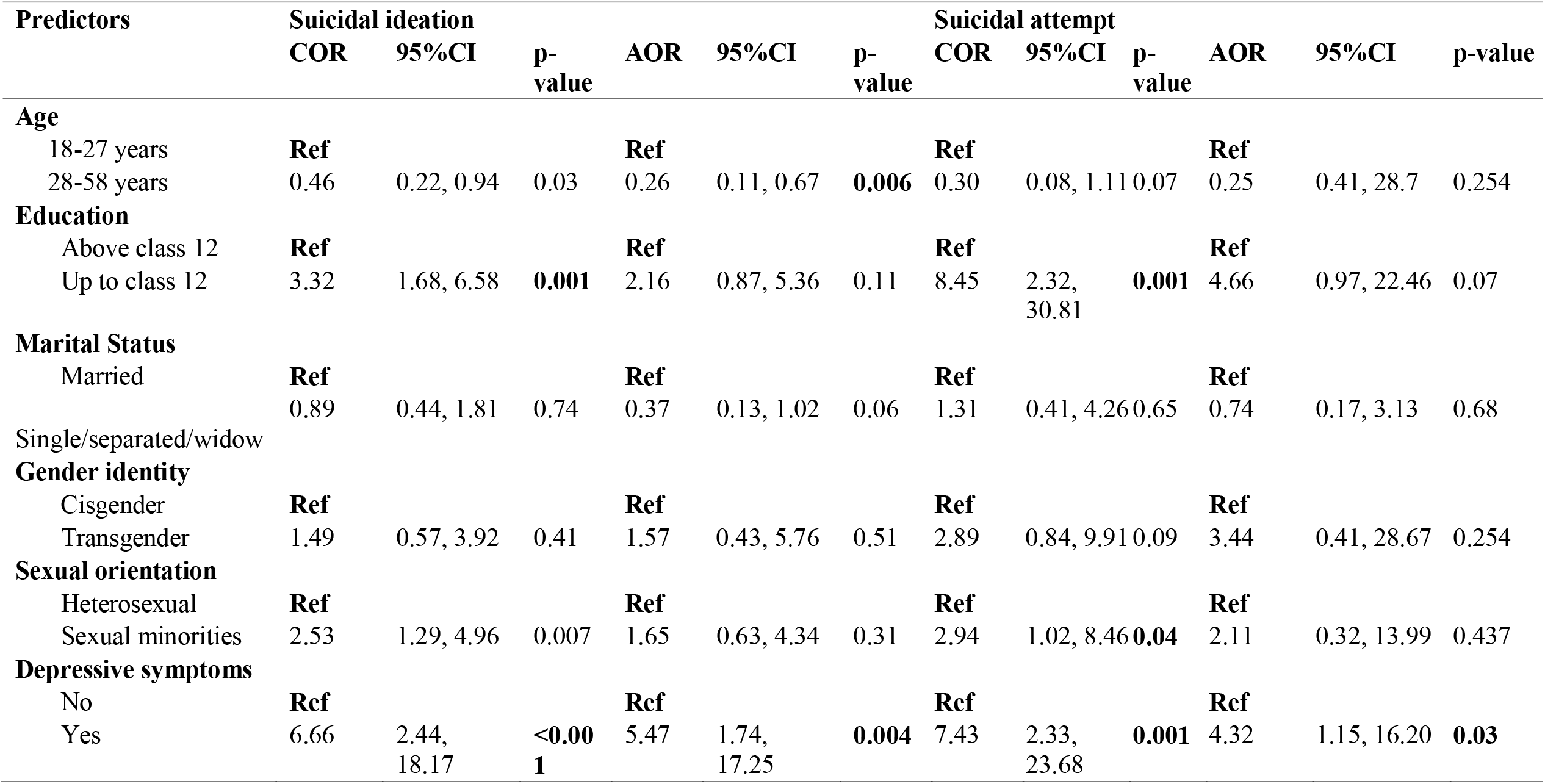
Factors associated with suicidal ideation and attempt (logistic regression) (n=190)

### Qualitative findings

IDIs of five participants from SGM group and one from heterosexual cisgender group who reported attempting suicide in last six months and four participants from heterosexual cisgender group experiencing depressive symptoms were taken. All participants were asked to share about their lived experiences of suicidal attempts, depressive symptoms or both.

### Theme 1: Depressive symptoms and its awareness

Although most of the participants were well aware about having and experiencing depressive symptoms, some of them mentioned that it might be normal to experience such symptoms and perhaps everyone these days feels the same. For example, one of the non-SGM participants said,

> ***“These days, I think everyone is having depressive symptoms and I feel like it is normal to have it. I assume each person has some mental health issues that they are facing, so I think it is normal in this era to have depressive symptoms*.*”***

Most commonly experienced symptoms of depression as stated by participants were not willing to talk to anyone, not preferring social interactions, difficulty to sleep at night or feeling excessively sleepy/tired during day and loss of appetite. One participant mentioned that they do not even feel like attending classes and participating in group activities.

### Theme 2: Factors leading to depressive symptoms/suicidal attempts

Participants stated that there were not one but combination of factors leading to either depression or suicidal attempts. Some of the factors commonly stated were financial challenges, relationship problems, familial crisis, stress due to exams, loneliness, loss of close friends and lack of family support during those periods of depressive episodes and suicidal attempts. One participant also mentioned that the college environment for them was not favourable or friendly to establish close relationships with their friends. Another participant mentioned that they were sexually abused as a child and it has been traumatising thus, leading to mental health problems.

Example of familial crisis include, family restrictions and evil behaviour of parents towards siblings for having romantic relationships beyond family’s permission, parental loss leading to regular maltreatment by other family members. Example of relationship problems include controlling behaviours of partner, break-ups with partner and partners not accepted by parents. One of the SGM participants stated, ***“…. they get angry very fast, yell at me for talking with my friends that they don’t prefer and sometimes even hit me for same reasons………”***

Only one out of five participants from SGMs said that social stigmas on their sexual orientation was one of the factors leading to depressive symptoms/suicidal attempts, the mediating cause being social exclusion among their friends. They stated, ***“… people used to bully me about***

> ***my sexual orientation, they tell me to wear feminine dresses because even being male I used to love hanging out with females a lot since my childhood, I loved playing with female friends, they used to call me “*Chhakka”, so I used to think it’s better to die than to live like this “***

(*Chhakka= commonly used word to verbally abuse or bully SGMs in Nepal.)

Factors leading to depressive symptoms/suicidal attempts in other four participants were not related to stigmas surrounding gender identity and sexual orientation, but stressors as mentioned previously. One of the SGM participants said, ***“…. maybe certain portion of population are affected because of homophobic/transphobic social stigmas, but not me; it has never made a difference to me on what people thought about my sexual orientation…*.*”***

### Theme 3: Motivating factors to relieve depressive symptoms/suicidal ideations

Self-motivation was one of the commonly stated motivating factors by the participants to overcome depressive symptoms/suicidal attempts, meaning that they themselves were the one who worked hard and did not give up on themselves and that was the most effective way to relieve depressive symptoms/suicidal ideations. They mentioned that getting busy with work was one of the self-motivating methods, the others were binge watching their favourite shows, chatting with family and friends, contemplating about being successful person in future, thinking about maintaining dignity of family. Some of them mentioned that their siblings and friends also encouraged them to overcome depressive symptoms/suicidal ideations by taking them out to visit new places, explore new food items, all such recreational activities. Participants mentioned that listening to their favourite music also helped deal with depressive episodes.

One of the SGM participants stated, ***“… I thought if I were to commit suicide, who will be the one to look after my mom? Will she be able to survive my suicide? And these thoughts also helped me overcome suicidal thoughts……”***

### Theme 4: Barriers to seek medical help

Only one out of ten participants said that they had consulted a doctor before for their depressive symptoms. Others have never sought medical help. As aforementioned, some even thought that it is not necessary to seek medical help because they thought everybody these days are having mental issues and they thought that it is normal to experience depressive symptoms or have suicidal thoughts. Some also mentioned that they are already having financial issues so, it is unreasonable for them to bear the cost of doctor’s fees, counselling charge and cost of medications, to overcome mental health issues.

One of them also mentioned that they are afraid once they take medicines, they might have to take it life long and get addicted to those medicines. Thus, not wanting to depend on medicines was also one of the reasons for not seeking medical help. They stated, ***“… what if I went to the doctor, he prescribed me with medicines and I have to continue it forever, and my sister also counselled me that it is better to die than to visit doctor for regular psychiatric medicines………”***

### Theme 5: Support from family/friends

Only two out of ten participants said that they had strong family/friends support that they could receive when in need. One of them shared their problems with their sister while the other had a close friend outside family to rely on during vulnerable times. Other eight participants had several reasons not to share their problems with their friends and family. They said that people did not take their problems seriously, that they were ignored a lot by friends and family even when they attempted to share. One of the SGM participants stated, ***“…***.. ***I think everyone is only worried about themselves, no one cares about what the other person is feeling, so it is better to keep everything to myself than to get humiliated by putting myself out there in front of other people……”***

Another from non-SGM group said, ***“…… I cannot share it with my family members because they will say that it’s all drama……”***

## Discussion

Overall, the findings of this study revealed that 10% of the participants had depressive symptoms with the mean score being 4.7±3.9. The prevalence of depressive symptoms in gender minorities was 13.6% with the mean score of 6.1±4.6. This is in contrast with the findings from a cohort study done in the U.S. where there were 52% depressive symptoms prevalent in gender minorities with the mean score of 10.7 on 30 point likert scale (CESD-10).^16^ The mean age of the sample is comparable, being 27.9 years for this study while 25.7 years for the study in the U.S,^16^ hence age cannot be the factor for such discrepancy in prevalence of depression. Therefore, the reason for such lower prevalence in our study might be because of the fact that those who participated in our study were somehow in contact with the organizations working in support of the SGM people. These organizations mentioned periodic screening of SGM carried out by them regarding depression and also referring them to health care institutions whenever necessary.

They also mentioned about the availability of the mental health counsellor in their workplace who counselled people when required. Another study showed that the prevalence of depressive symptoms in gender minorities was 33.3%.^17^ Thus, we can say that among gender minorities, depressive symptoms scores seem to be highly variable.

Likewise, the mean score of depressive symptoms for sexual minorities in our study was significantly higher (6.3±4) compared to heterosexual participants (3.8±3.6). This is in line with the study conducted in the Finnish population where sexual minority participants had significantly higher mean depressive scores (14.2±0.19) compared to their heterosexual counterparts (11.9±0.06).^8^ This can perhaps be explained by the presence of chronic exposure of sexual minorities to stigma, victimization, identity concealment, and lower family support, attributed to minority stress model.

The prevalence of suicidal attempts was 18.2% in gender minorities which is higher than in cisgender population (7.1%). This is comparatively lower with the findings of a study conducted in Canada which showed 36.5% lifetime prevalence of suicidal attempts in gender minorities compared to three percent in heterosexual people.^18^ Further, our study showed 13.7% lifetime suicidal attempts prevalence in sexual minorities compared to 5.1% in heterosexual people. This is in line with the study of Canada where lifetime prevalence of suicidal attempts in sexual minorities was 13.1% and that in heterosexual people was only 2.8%.^18^

Similarly, average depressive symptoms scores of gender minorities were significantly higher (β= 2.47, p=0.009) compared to cisgender participants, in model 1 while average score of sexual minorities were also significantly higher (β= 2.47, p<0.001) as compared to heterosexual participants. One of the studies done in the U.S. has also showed that cisgender females (β= 0.21, p<0.01) and transgender males/females (β= 0.66, p<0.01) reported higher depressive symptoms than the cisgender males.^19^ Comparing these findings with another study conducted in Taiwan, it is of our interest that the study did not show significant association either with gender identity or with sexual orientation but it showed significant association of higher familial sexual stigma (β= 0.25, p<0.001), higher internalized sexual stigma (β= 0.15, p<0.001) and lower perceived family support (β= -0.87, p<0.001) with higher depressive symptoms.^20^ These results suggest that the mental health issues like depressive symptoms are not merely associated with the sexual orientation or the gender identity of individuals, rather the social and familial stigmas related to homophobia and transphobia are the root causes of such issues. Furthermore, our study did not show significant association of age with depressive symptoms. This conflicts with other studies revealing association with younger age in transgender people (p<0.01).^17^ Shared experiences of stigma, and social exclusion across all ages might have overshadowed the age-related differences in depressive symptoms in Nepali SGM communities.

Moreover, model 2 showed higher depressive symptoms (β= 2.11, p=0.006) in victims of bullying related to SOGI which is comparable with the results of study done in Latinx SGM youth (β= 0.20, p<0.01).^19^ Our study also showed significant association of single/separated/widowed participants (β= 1.40, p=0.036) with higher depressive symptoms compared to married ones in model 1 which is similar to results of a study in rural China (OR= 1.39, p<0.001).^21^ However, it also suggests that significant mediating effects of sleeping time, existing pain and life satisfaction of an individual on the correlation between marital status and depression (p<0.001).^21^ Despite Nepal’s 2023 step toward marriage equality with provisions for temporary registration for SGM couples, social acceptance remains low with many still unmarried while others are forced into marriages outside their will.

Our study showed that the participants with depressive symptoms had 5.47 times (95% CI: 1.74, 17.25) higher odds of having suicidal ideations which is in line with Tegegne’s study (OR: 4.88).^22^ Another study done in Korea also support this result.^23^ Moreover, it also suggests mediating effects of social support in between depression and suicidal ideation.^23^ Further, the findings showed that a significant association between suicidal attempts history and increased depressive symptoms (AOR: 4.32, 95% CI: 1.15, 16.20, p=0.03) which is supported by a study done in Malaysia (AOR: 4.3, 95%CI: 3.9, 4.8).^24^ Our findings also showed that the older participants (28-58 years) were 74 times (AOR= 0.26, 95% CI: 0.11, 0.67, p=0.006) less likely to have suicidal ideations which is supported by a study in Malaysia.^25^

Qualitative analysis showed that there were combination of common factors leading to depressive symptoms /suicidal attempts in SGM population as well as in heterosexual cisgender population. They were financial hardships, relationships problems, familial crises, educational stress, lack of close friends and family support. One of the reasons participants mentioned in the in-depth interviews for not sharing how they feel with others, was because they were afraid that people would not believe them and they would think that it is all dramatic. They said that others did not take their problems seriously and that they were ignored a lot by closed ones even when they attempted to share. This appeared consistent with previous research which stated that help-seeking for suicidal distress has been stigmatised in our society and it has been re-casted as attention seeking rather than help seeking.^26^ Thus, even when people are in dire need of social support from their family and friends during severe depressive episodes or during suicidal attempts, others can misunderstand it as an attention seeking behaviour.^27^ Perhaps this is also one of the reasons that participants were struggling on their own and downplaying suicidal thoughts and depressive symptoms. They even thought that all these experiences are normal and possibly everyone these days feels the same. This can also be correlated with the fact that only one out of ten participants had ever sought medical help for their depressive symptoms and suicidal thoughts. If these misunderstandings between the sufferer and the society persist then it might lead to an eternal cycle of depressive symptoms for people especially among marginalized groups like SGM individuals.

One of the major limitations of this study is that it could not capture people who have died by suicidal attempts, studies on actual suicidal counts is recommended by approaching the family members and closed ones of people who committed suicide and died in order to receive more accurate findings.

## Conclusions

The depressive symptoms as well as suicidal attempts were higher among sexual and gender minorities than in heterosexual cisgender people. However, depressive symptoms and suicidal attempts were mostly led by social stigmas related to mental health issues and SGMs, rather than individual’s own gender identity or sexual orientation. Moreover, the depressive symptoms were highly related with the bullying and this was further ascertained during in-depth interviews.

Therefore, it is recommended that further research be done on the social stigmas and its impacts on SGMs’ mental health.

### Clinical implications

Suitable environment for SGM should be fostered in health care facility to improve their health-seeking behaviour. Bullying based on SOGI should be prohibited in the community to decrease depression and suicide in SGM. Interventions to reduce social stigmas related to SOGI and mental health issues is paramount.

## Data Availability

All data produced in the present study are available upon reasonable request to the authors

## List of abbreviations

IDI: In-depth Interview
LGBTI: Lesbian, Gay, Bisexual, Transgender, Intersex
SDG: Sustainable development goal
SGM: Sexual and gender minority
SOGI: Sexual orientation and gender identity
SOGIESC: Sexual orientation, gender identity and expression, and sex characteristics
TGD: Trans and gender diverse

## Declarations

### Ethics approval and consent to participate

The ethical approval was obtained from Institutional Review Committee (IRC) of Institute of medicine, Tribhuvan University (Ref. no. 358 (6-11) E2). Informed consent was taken from all participants. Privacy and confidentiality were maintained as much as possible. Those participants found to be reporting severe depression and not seeking professional help were suggested to seek help and were provided with national hotline numbers for suicide prevention. This study adheres to the principles of the Declaration of Helsinki.

### Consent for publication

Not applicable

### Availability of data and materials

The datasets used and/or analysed during the current study are available from the corresponding author on reasonable request.

## Author’s contributions

BS and BPC designed and conceptualized the study. BS and MPK had a major role in data acquisition. BS, BP and RB were involved in data analysis and interpretation. All authors were major contributors to writing the manuscript. All the authors rigorously contributed in editing, revising and proofreading the manuscript. All the authors read and approved the final manuscript.

## Acknowledgements

Not applicable

## References

1. Asian Development Bank. Summary of the Analytical Study for the Safeguard Policy Review and Update: Sexual Orientation, Gender Identity, Gender Expression, and Sex Characteristics (SOGIESC)[Internet]. Asian Development Bank; 2025 May [cited 2025 Aug 8]. Available from: https://www.adb.org/documents/spru-analytical-study-summary-sogiesc-draft

2. ARC International. Yogyakarta Principles plus 10 [Internet]. Geneva: ARC International; 2017 Nov 10 [cited 2025 Aug 8]. Available from: https://yogyakartaprinciples.org/principles-en/yp10/

3. Constituent Assembly Secretariat. Constitution of Nepal 2015 [Internet]. Kathmandu: Constituent Assembly Secretariat; 2015 [cited 2025 Aug 9]. Available from: https://ag.gov.np/files/Constitution-of-Nepal_2072_Eng_www.moljpa.gov_.npDate-72_11_16.pdf

4. Human Rights Watch. We Have to Beg So Many People [Internet]. New York: Human Rights Watch; 2024 Feb 15 [cited 2025 Aug 9]. Available from: https://www.hrw.org/report/2024/02/15/we-have-beg-so-many-people/human-rights-violations-nepals-legal-gender

5. UN Women. Addressing violence against LGBTIQ+ people in Nepal [Internet]. UN Women; 2023 June 11 [cited 2025 Aug 9]. Available from: https://un.org.np/sites/default/files/doc_publication/2023-06/LGBTIQ%20Study%20Report-Final-web%20version-11%20June%202023%20evening.pdf

6. WHO. Improving LGBTIQ+ health and well-being with consideration for SOGIESC [Internet]. WHO; [cited 2025 Aug 9]. Available from: https://www.who.int/activities/improving-lgbtqi-health-and-well--being-with-consideration-for-sogiesc

7. UCLA School of Law Williams Institute. Surveying Nepal’s Sexual and Gender Minorities. 2014 Oct [cited 2023 Mar 17]. Available from: https://williamsinstitute.law.ucla.edu/publications/survey-sgm-nepal/

8. Harper GW, Crawford J, Lewis K, Mwochi CR, Johnson G, Okoth C, et al. Mental Health Challenges and Needs among Sexual and Gender Minority People in Western Kenya. International Journal of Environmental Research and Public Health [Internet]. 2021 Jan [cited 2023 Mar 17];18(3):1311. Available from: https://www.mdpi.com/1660-4601/18/3/1311

9. KaLJllstroLJm M, Nousiainen N, Jern P, Nickull S, Gunst A. Mental health among sexual and gender minorities: A Finnish population-based study of anxiety and depression discrepancies between individuals of diverse sexual orientations and gender minorities and the majority population. PLOS ONE [Internet]. 2022 Nov 4 [cited 2023 Mar 17];17(11):e0276550. Available from: https://journals.plos.org/plosone/article?id=10.1371/journal.pone.0276550

10. Gmelin JOH, De Vries YA, Baams L, Aguilar-Gaxiola S, Alonso J, Borges G, et al. Increased risks for mental disorders among LGB individuals: cross-national evidence from the World Mental Health Surveys. Soc Psychiatry Psychiatr Epidemiol. 2022 Nov 1;57(11):2319–32.

11. Olson KR, Kuper LE, Wittlin NM. Mental Health of Transgender and Gender Diverse Youth. Annu Rev Clin Psychol [Internet]. 2023 Jan 6;19:207-232. Available from: https://pmc.ncbi.nlm.nih.gov/articles/PMC9936952/

12. Surace T, Fusar-Poli L, Vozza L, Cavone V, Arcidiacono C, Mammano R, et al. Lifetime prevalence of suicidal ideation and suicidal behaviors in gender non-conforming youths: a meta-analysis. Eur Child Adolesc Psychiatry. 2021 Aug;30(8):1147–61.

13. Argüello TM. Decriminalizing LGBTQ+: Reproducing and resisting mental health inequities. CNS Spectr. 2020 Oct;25(5):667–86.

14. United Nations. Transforming our world: The 2030 Agenda for Sustainable Development [Internet]. [cited 2023 Mar 17]. Available from: https://sustainabledevelopment.un.org/content/documents/21252030%20Agenda%20for%20Sustainable%20Development%20web.pdf

15. Kohrt BA, Luitel NP, Acharya P, Jordans MJD. Detection of depression in low resource settings: validation of the Patient Health Questionnaire (PHQ-9) and cultural concepts of distress in Nepal. BMC Psychiatry. 2016 Dec;16(1):58.

16. Reisner SL, Katz-Wise SL, Gordon AR, Corliss HL, Austin SB. Social epidemiology of depression and anxiety by gender identity. J Adolesc Health. 2016 Aug;59(2):203– 8.

17. Hajek A, KoLJnig HH, Buczak-Stec E, Blessmann M, Grupp K. Prevalence and Determinants of Depressive and Anxiety Symptoms among Transgender People: Results of a Survey. Healthcare (Basel). 2023 Feb 27;11(5):705.

18. Liu L, Batomen B, Pollock NJ, Contreras G, Jackson B, Pan S, et al. Suicidality and protective factors among sexual and gender minority youth and adults in Canada: a cross-sectional, population-based study. BMC Public Health. 2023 Aug 2;23(1):1469.

19. Abreu RL, Lefevor GT, Gonzalez KA, Barrita AM, Watson RJ. Bullying, depression, and parental acceptance in a sample of Latinx sexual and gender minority youth. Journal of LGBT Youth. 2023 Jul 3;20(3):585–602.

20. Wang PW, Chang YP, Tsai CS, Yen CF. Predictors of depressive and anxiety symptoms among lesbian, gay and bisexual young adult individuals experiencing the COVID-19 pandemic: A four-year follow-up study. Journal of the Formosan Medical Association [Internet]. 2024 Feb 28 [cited 2024 Mar 10]; Available from: https://www.sciencedirect.com/science/article/pii/S0929664624001384

21. Pan L, Li L, Peng H, Fan L, Liao J, Wang M, et al. Association of depressive symptoms with marital status among the middle-aged and elderly in Rural China– Serial mediating effects of sleep time, pain and life satisfaction. Journal of Affective Disorders. 2022 Apr 15;303:52–7.

22. Tegegne K, Tesfaye E, Tessema M, Bagajjo W, Dumo M, Abebe A, et al. The Association between Depression and Suicidal Ideation A Systematic Review and Meta-Analysis. Psychology and Mental Health Care. 2022 Jan 5;6:01–6.

23. Kim BJ, Kihl T. Suicidal ideation associated with depression and social support: a survey-based analysis of older adults in South Korea. BMC Psychiatry. 2021 Dec;21(1):1–9.

24. Aziz FAA, Razak MAA, Ahmad NA, Awaluddin SM, Lodz NA, Sooryanarayana R, et al. Factors Associated With Suicidal Attempt Among School-Going Adolescents in Malaysia. Asia Pacific Journal of Public Health. 2019;31(8):73S–79S.

25. Cheah YK, Azahadi M, Phang SN, Abd Manaf NH. Association of Suicidal Ideation with Demographic, Lifestyle and Health Factors in Malaysians. East Asian Arch Psychiatry. 2018 Sep;28(3):85–94.

26. McDermott E, Hughes E, Rawlings V. Norms and normalisation: understanding lesbian, gay, bisexual, transgender and queer youth, suicidality and help-seeking. Culture, Health & Sexuality. 2018 Feb 1;20(2):156–72.

27. Chandler A. Self-injury, medicine and society: Authentic bodies. New York, NY: Palgrave Macmillan/Springer Nature; 2016. xii, 217 p. (Self-injury, medicine and society: Authentic bodies).

